# *Streptococcus intermedius* as a leading agent of brain abscess: retrospective analysis of a UK cohort

**DOI:** 10.1101/2020.01.25.20018788

**Authors:** Christopher A Darlow, Nicholas McGlashan, Richard Kerr, Sarah Oakley, Pieter Pretorius, Nicola Jones, Philippa C Matthews

## Abstract

**Background:** Brain abscess is an uncommon condition, but carries high mortality. Current treatment guidelines are based on limited data. Surveillance of clinical, radiological and microbiology data is important to inform patient stratification, interventions, and antimicrobial stewardship.

**Methods:** We undertook a retrospective, observational study of patients with brain abscess, based on hospital coding, in a UK tertiary referral teaching hospital. We reviewed imaging data, laboratory microbiology, and antibiotic prescriptions.

**Results:** Over a 47 month period, we identified 47 adults with bacterial brain abscess (77% male, median age 47 years). Most of the abscesses were solitary frontal or parietal lesions. A microbiological diagnosis was secured in 39/47 (83%) of cases, among which the majority were of the *Streptococcus milleri* group (27/39; 69%), with a predominance of *Streptococcus intermedius* (19/27; 70%). Patients received a median of 6 weeks of intravenous antibiotics (most commonly ceftriaxone), followed by variable oral follow-on regimens. Ten patients (21%) died, up to 146 days after diagnosis. Mortality was significantly associated with increasing age, multiple abscesses, immunosuppression and the presence of an underlying cardiac anomaly.

**Conclusion:** Our data suggest that there has been a shift away from staphylococcal brain abscesses, towards *S. intermedius* as a dominant pathogen. In our setting, empiric current first line therapy with ceftriaxone remains appropriate on microbiological grounds and narrower spectrum therapy may sometimes be justified. Mortality of this condition remains high among patients with comorbidity. Prospective studies are required to inform optimum dose, route and duration of antimicrobial therapy.

## INTRODUCTION

Brain abscesses are a focal infection characterised by a walled-off collection of pus within the brain parenchyma. They may arise spontaneously, or as a result of specific risk factors including intravenous drug use, congenital cardiac defects, infective endocarditis, immunosuppression predisposing to infection, or a contiguous focus of infection spreading directly to the adjacent central nervous system (CNS), for example from dental, sinus, middle or inner ear infection (1–5). Previous reports suggest that brain abscesses predominantly occur in males (accounting for around 70% of cases) and affect young adults (6,7).

Brain abscesses are usually caused by bacteria, although can also be caused by other pathogens including fungi and protozoa (4). Prior to 1960, staphylococcal aetiology had been the commonest cause, accounting for over a third of cases, with streptococcal causes accounting for around a further 30% (6). However, in recent decades, the incidence of staphylococcal brain abscesses has been falling, with streptococci increasingly dominating (6,7). Anaerobes are also described as important contributors, and polymicrobial aetiology is recognised in around a quarter of cases, although this may be underestimated according to the limited sensitivity of laboratory diagnosis. However, evidence is limited, as studies before 2010 report identification of a pathogen in only approximately 60% of cases (8,9). With modern stereotactic neurosurgical sampling methods and advances in molecular diagnostics, there is scope for improving rates of identification of specific pathogens, up to ∼80% in the more recent literature, (7) providing an opportunity to review approaches to antimicrobial therapy.

There are no recent unifying international or national guidelines for the management of bacterial brain abscesses. An ‘Infection in Neurosurgery Working Party’ published antibiotic recommendations on behalf of the British Society of Antimicrobial Chemotherapy in 2000 (10). However, these recommendations were based on very limited published data, and are not universally adopted in clinical practice. Other recommendations are made elsewhere in the existing literature, including the need to rule out other causes of space-occupying lesions (such as primary or metastatic malignancy), and additional investigations including an HIV test and consideration of cardiac echocardiography (4,5,11). The recommended empirical antimicrobial treatment for brain abscesses is often intravenous ceftriaxone or cefotaxime (5,12), which cover the likely gram-positive causes, as well as providing coverage for the less common gram negative organisms, and offering sufficient central nervous system penetration. Adjunctive metronidazole may also be recommended to cover anaerobic organisms (10). However, the suggested choice, duration and route of antibiotic therapy is based on scarce evidence (13). In the current era of increasing antimicrobial resistance, careful scrutiny of laboratory data is required to ensure that empiric treatment regimens are sufficiently broad to cover the majority of likely pathogens, while at the same time limiting unnecessary exposure to broad-spectrum agents. Furthermore, new evidence for endocarditis and for musculoskeletal infection suggests that oral antibiotics are non-inferior to parenteral therapy in some deep-seated infections (14,15). At present there are no such data to inform the management of CNS infection, but the route of therapy remains an important question to be addressed.

Given this changing landscape of antimicrobial therapy, the paucity of epidemiological data, and the limited guidelines for brain abscesses, we set out to collect clinical and laboratory data from a large UK teaching hospital to examine the epidemiological and microbiological trends of bacterial brain abscesses. We specifically undertook to address: i) Any evidence for changes in the epidemiology or microbiology of brain abscesses over time; ii) The extent to which current antimicrobial management strategies are appropriate to cover the organisms identified, considering both empirical and directed therapy; iii) Determination of any host factors that are associated with specific outcomes, in order to identify high risk cases, with implications for prognosis, monitoring and interventions.

## METHODS

### Data Collection

We collected data retrospectively from the electronic patient records of Oxford University Hospitals (OUH) NHS Foundation Trust, a large teaching hospital trust in the Thames Valley region of the UK and a tertiary referral centre for neurosurgery. We identified patients using coding data (any patients coded with ‘G06.0’, the ICD code for ‘Intracranial abscess and granuloma’) admitted between 1^st^ January 2013 and 1^st^ December 2016. We screened individual records to remove paediatric cases, erroneously coded cases and those with non-bacterial causes (e.g. *Toxoplasma gondii*), and then reviewed electronic notes and results, extracting the relevant data. The study was undertaken as a registered audit to determine the quality of management of brain abscesses. Approval for this was given internally via the OUH audit management team and we collected and stored data in accordance with relevant governance standards. All data were anonymised prior to analysis.

We audited each case for adherence to local antimicrobial guidelines, which are accessible to clinical staff via intranet and the smartphone app ‘Microguide’. For bacterial brain abscesses, the antimicrobial advice is as follows:

- Urgent biopsy with collection of specimens for culture is recommended in most cases;
- Empiric choice of antimicrobial therapy is ceftriaxone (2g iv bd) and metronidazole (400mg tds po);
- Substitute the cephalosporin with ciprofloxacin in patients with a history of beta-lactam allergy;
- Add vancomycin in cases known to be colonised or infected with meticillin resistant *Staphylococcus aureus*;
- For patients at risk of immunosuppression, consider amending empiric therapy to broaden spectrum of cover;
- Modify initial treatment according to culture and susceptibility data;
- Consult a microbiology/infectious diseases team for clinical review and advice;
- Treatment is recommended for 4-8 weeks.

### Radiology data

Brain abscesses in our cohort were typically diagnosed by computer topography (CT) scan, in some cases followed by magnetic resonance imaging (MRI) to delineate further. We collected radiology data for the definitive pre-treatment scan only, as subsequent sequential scans varied according to clinical need. For the purposes of data collection for our study, all scans were retrospectively reviewed by two independent radiologists, blinded to all other clinical and microbiological data. To calculate the size of an abscess in cross-section, we assumed an elliptical area (area (mm^3^) = 0.5 × maximum width (mm) × 0.5 × maximum length (mm) × π).

### Laboratory diagnosis

Microbiology diagnostic work was undertaken in our ISO approved clinical diagnostic laboratory. Methods were based on National Standard Methods (PHE) (16). In brief, samples from brain abscesses were handled as follows: a Gram stain was undertaken and the sample was inoculated into (i) cooked meat enrichment broth (extended incubation for 10 days in aerobic conditions), (ii) blood, chocolate and MacKonkey agar incubated in 5% CO2, (iii) blood, NAT and neomycin agar (in anaerobic conditions with metronidazole discs). All were incubated at 35-37°C. Growth on any plate was followed up by identification using by mass spectrometry (Maldi-TOF). Antibiotic susceptibility was determined on the automated Phoenix platform (Becton Dickinson). Examination for additional specific pathogens would be undertaken at the request of the clinical team or based on attributes of the patient’s background or presenting features (e.g. mycobacteria, nocardia, fungi, acanthamoeba).

Blood cultures were inoculated into BACTEC bottles under aseptic technique and incubated on the Becton Dickinson BACTEC FX system, using a standard operating procedure based on SMI B 37: ‘investigation of blood cultures’ (17).

### Data regarding surgical and medical management

A limited dataset regarding surgical intervention and antimicrobial therapy was collected from the electronic patient record. Antimicrobial therapy was monitored and modified under supervision of the Infectious Diseases / Microbiology team according to the clinical context and microbiology results. For patients well enough to be discharged but requiring ongoing parenteral therapy, the full course of antibiotics was administered under the supervision of the Oxford outpatient antimicrobial therapy (OPAT) team via a peripherally inserted central catheter (PICC) (18).

### Statistical methods

Descriptive statistics of the data was collated using Microsoft Excel and SPSS software packages. Univariate associations were explored using Fisher’s Exact Test, one-way ANOVA or Kruskal-Wallis statistical tests, depending on the modality and distribution of the data.

## RESULTS

### Brain abscess patients presented to our service at an average rate of 1 per month and were predominantly male

Over the study time period of 47 months, 74 patients were coded by the hospital as G06.0. Of these, 47 were aged ≥16 years and had a final diagnosis of a bacterial brain abscess (Table 1; Suppl Fig 1). All the data reported in this study are provided as a metadata table in xls format (see link in supplementary data section). The majority of our cases were male (n=36; 77%), in keeping with previous case series (8,9). The median age was 47 years (range 17-91). An identifiable risk factor for brain abscesses was identified in 26 (55%; Table 1).

**Table 1:** Demographic and host characteristics of a brain abscess cohort of 47 adults recruited in a UK tertiary referral hospital in the UK. sd=standard deviation; IQR=inter-quartile range.

| Demographics | Male | Female | Total |
| --- | --- | --- | --- |
| Total number of patients (%) | 36 (76.6) | 11 (23.4) | 47 (100) |
| Median age at presentation (sd) | 46.6 (15.8) | 63.0 (20.5) | 46.9 (16.9) |
| Range of age at presentation (years)<br>(IQR) | 17 – 89<br>(38 - 59) | 23 – 91<br>(41– 64) | 17 – 91<br>(38 – 61) |
| Mortality (%) | 6/36 (16.7) | 4/11 (36.4) | 10/47 (21.3) |
| **Risk Factors\*:** |  |  |  |
| Intravenous Drug Use (%) | 5/36 (13.9) | 2/11 (18.2) | 7/47 (14.9) |
| Immunosuppression** (%) | 1/36 (2.8) | 1/11 (9.1) | 2/47 (4.6) |
| Cardiac Anomaly (%) | 7/36 (19.4) | 1/11 (9.1) | 8/47 (17.0) |
| Dental / ENT source (%) | 10/36 (27.8) | 4/11 (36.4) | 14/47 (29.8) |
| No risk factor identified (%) | 17/36 (47.2) | 4/11 (36.4) | 21/47 (44.7) |

\*Some individuals have > 1 risk factor(s).
\*\*No patients were HIV positive

### Brain abscesses were typically single lesions caused by Streptococcus intermedius

The majority of abscesses were a single lesion (n=36; 77%), commonest sites were frontal (n=15; 32%) and parietal (n=8; 17%); Table 2. Microbiological diagnosis was made in 39/47 cases (83%; Table 3), of which 34 were cultured from pus and five from blood cultures. Strikingly, organisms of the *S. milleri* group accounted for 29/39 cases in which diagnostic data were available (74%), among which *S. intermedius* accounted for the majority (19/29; 66%). In one case, *S. milleri* was identified from an orbital swab that had been collected in another clinical centre. Although the result of a swab should be interpreted with caution as potentially reflecting only colonising or commensal flora, in this case the growth of *S. milleri* is a plausible agent of deep-seated infection.

**Table 2:** Radiological features of brain abscesses at time of presentation.

| Features of abscess | Number (%) |
| --- | --- |
| **Number of abscesses** |  |
| Single | 36/47 (77%) |
| Multiple | 11/47 (23%) |
| **Size of abscess** |  |
| Median cross-sectional size of abscess, mm<sup>2</sup> (IQR) | 500 (199-889) |
| **Location of abscess(es):** |  |
| Frontal | 15/47 (32%) |
| Parietal | 8/47 (17%) |
| Temporal | 7/47 (15%) |
| Occipital | 4/47 (9%) |
| Cerebellum | 2/47 (4%) |
| Other subcortical location | 2/47 (4%) |
| Multiple locations | 9/47 (19%) |
| **Radiological features:** |  |
| Abscess wall enhancement | 42/44* (95%) |
| Dual Rim Sign present | 10/44* (23%) |
| Oedema present in one lobe | 31/47 (66%) |
| Oedema present in >1 lobe | 15/47 (32%) |
| Ventriculitis present | 5/47 (11%) |
| Sinus involvement | 7/47 (15%) |

\*Difference in denominator due to three patients not receiving contrast enhancement

**Table 3:** Pathogens identified in a cohort of 47 adults with a diagnosis of brain abscess.

| Causative Organism | Number of cases (%) | Ceftriaxone Sensitivity |
| --- | --- | --- |
| **Gram positive** |  |  |
| *Streptococcus intermedius*<sup>a,b</sup> | 20 (43%) | 20/20 (100%) |
| *Streptococcus constellatus*<sup>a,b</sup> | 4 (9%) | 4/4 (100%) |
| *Streptococcus anginosus*<sup>a</sup> | 4 (8.5%) | 4/4 (100%) |
| *Streptococcus milleri* group (not further identified)<sup>c</sup> | 1 (17.0%) | 1/1 (100%) |
| *Staphylococcus aureus* | 1 (2.1%) | 1/1 (100%) |
| *Listeria monocytogenes*<sup>e</sup> | 2 (4.3%) | 0/2 (0%) |
| **Gram negative or mixed** |  |  |
| *E. coli* | 1 (2.1%) | 1/1 (100%) |
| *Pseudomonas aeruginosa* | 1 (2.1%) | 0/1 (0%) |
| *Fusobacterium nucleatum*<sup>f</sup> | 3 (6.4%) | N/A |
| *Citrobacter*, *Pseudomonas*, *Corynebacterium* + anaerobes | 1 (2.1%) | 0/1 (0%) |
| *Aggregatibacter aphrophilus* + *Actinomyces meyeri* | 1 (2.1%) | 1/1 (100%) |
| **No organism identified** |  |  |
| Sterile sample | 4 (8.5%) | N/A |
| No sample taken | 4 (8.5%) | N/A |

<sup>a</sup> These organisms are all part of the *S. milleri* group
<sup>b</sup> One *S. intermedius* and one *S. constellatus* reported in mixed culture with anaerobes
<sup>c</sup> Isolate grown from an orbital swab
<sup>d</sup> *S. aureus* sensitive to meticillin
<sup>e</sup> Both patients with *Listeria* infection were >55 years of age but neither had any known cause of immunosuppression
<sup>f</sup> Of the three cases with anaerobes isolated as a sole causative organism, two patients were intravenous drug users and the third had no clear identified source.

We investigated whether the presence of *S. milleri* was associated with any clinical or radiological characteristics and identified a significant relationship only with the size of the abscess (median cross-sectional size of abscess 930mm^3^ for *S. milleri* vs 190mm^3^ for other organisms; p=0.0005; Suppl Table 1).

### Neurosurgical drainage was undertaken in the majority of cases, followed by a median of six weeks intra-venous antibiotics

Among our 47 patients, 38 (81%) had an intervention to drain the abscess, and 15 underwent >1 drainage procedure (Suppl Table 2). The median time between admission for the brain abscess and first procedure (where first microbiological samples were taken) was 1 day (range 0 – 10 days). One individual had an elective biopsy for a presumed tumour. When the microbiology and histology suggested a diagnosis of a brain abscess, the patient was admitted for treatment 11 days later. All patients were treated with intravenous antibiotics, predominantly ceftriaxone, with alternative choices justified on the grounds of clinical context and/or microbiology (Table 4). Duration of IV therapy was most commonly 6 weeks while oral therapy varied from 2-8 weeks (Suppl Fig 2).

**Table 4:**
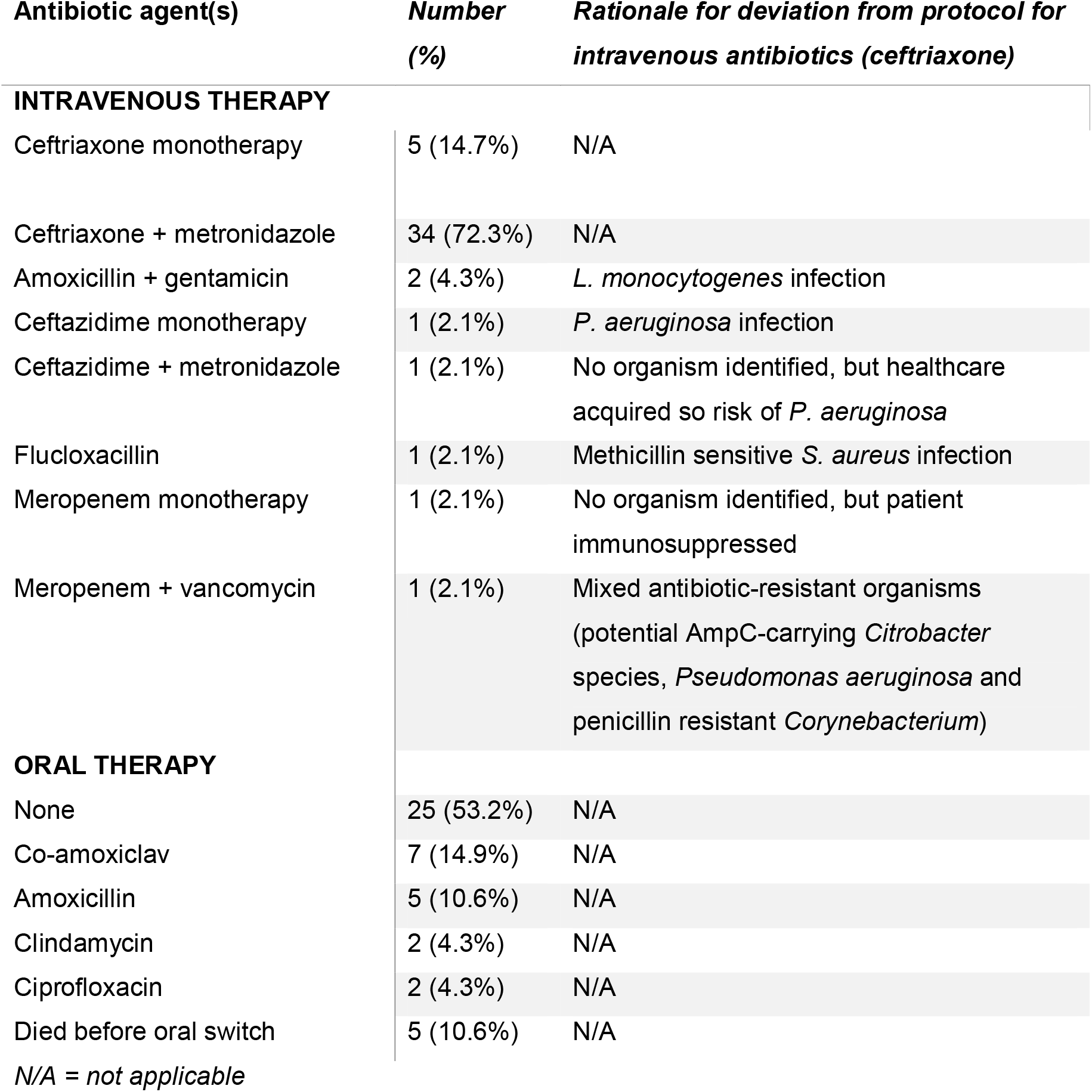
Intravenous and oral antibiotic regimens used to treat 46 adults with bacterial brain abscess. Data missing for one patient in cohort of 47.

### Brain abscesses are associated with high mortality

Ten patients died (21%). Five (50%) deaths occurred before completion of the primary intravenous antimicrobial course and were directly attributable to the brain abscess (median 9 days after presentation; range 1-50 days), three were after the completion of treatment but showed possible attribution to brain abscess (median time post-presentation 116 days; range 76-146 days). One was attributed to unrelated comorbidities (113 days post-presentation), and one died at another centre with no further data available (146 days post-presentation). Of the 10 deaths, seven had significant medical comorbidities, including immunosuppression (n=2), cardiac disease (n=3), and other underlying medical conditions (n=5).

Increasing age, immunosuppression, and the presence of an underlying cardiac anomaly were significantly associated with mortality on univariate analysis (p=0.005, p=0.04, p=0.03, respectively; Table 5). Mortality was not significantly associated with any radiological features or with any specific microbiological diagnosis.

**Table 5:** Associations between clinical features and mortality in a cohort of 47 adults with bacterial brain abscess.

| Characteristic | Died<br>(n=10) | Survived<br>(n=37) | p-value<sup>a</sup><br>(univariate) |
| --- | --- | --- | --- |
| Median Age (IQR) | 60<br>(47 – 77) | 46<br>(38 – 61) | **0.005** |
| Sex (proportion male) | 6/10 (60%) | 30/37 (81%) | 0.16 |
| Intravenous Drug Use | 2/10 (20%) | 5/37 (14%) | 0.61 |
| Cardiac Anomaly | 4/10 (40%) | 4/37 (11%) | **0.03** |
| Immunosuppressed<sup>b</sup> | 2/10 (20%) | 0/37 (0%) | **0.04** |
| Dental/ENT Source | 3/10 (30%) | 11/37 (30%) | 0.99 |
| Any risk factor present | 8/10 (80%) | 18/37 (49%) | 0.15 |
| Mean number of brain abscesses | 1.89 | 1.35 | **<0.001** |
| ≥1 abscess present | 4/10 (40%) | 7/37 (19%) | 0.21 |
| Median cross-sectional size of abscess, mm<sup>2</sup> (IQR) | 707<br>(186-1272) | 478<br>(236-865) | 0.26 |
| Aspiration/drainage undertaken | 7/10 (70%) | 31/37 (84%) | 0.33 |
| More than one aspiration/drainage procedure performed | 1/10 (10%) | 15/37 (41%) | 0.13 |
| Presence of microbiological diagnosis | 7/10 (70%) | 32/37 (86%) | 0.22 |
| *Streptococcus milleri* infection | 6/10 (60%) | 23/37 (62%) | 1.0 |
| *Streptococcus intermedius* infection | 3/10 (30%) | 16/27 (60%) | 0.15 |
| Oral antibiotics received<sup>c</sup> | 0/5 (0%) | 19/37 (51%) | 0.05 |

<sup>a</sup> p-values for categorical variables calculated using Fisher's Exact test, for continuous variables using one-way ANOVA or Kruskal-Wallis, dependent on distribution of the data. Bold font indicates significant p value (<0.05)
<sup>b</sup> An HIV test was recorded for 30/47 cases (64%); all were negative. It is not clear from our data whether the remaining 17 were offered a test.
<sup>c</sup> Oral antibiotics refers to follow-on therapy after completion of a primary intra-venous course. Five patients died before completion of primary intravenous antibiotics.

## DISCUSSION

### Microbiology

In this UK cohort of brain abscess patients, we found a low prevalence of Staphylococcal infection and a predominance of Streptococcal species, in particular *S. intermedius* of the *S. milleri* group (which together includes *S. constellatus, S. anginosus* and *S. intermedius*). We identified a pathogen in 83% of cases, which is comparable to another recent UK case series (7) and compares favourably with two other data sets in which <60% of cases had a microbiological diagnosis (8,9). Improvements in sampling techniques, especially with relation to the timing of initial antibiotics, together with advances in microbiological laboratory practice (including the use of the MALDI-TOF) are likely reasons for the increased rates of laboratory diagnosis. Current expansion of metagenomic approaches to replace or supplement traditional phenotype-based methods (19,20) may underpin future improvements in identification rates.

The dominance of *S. milleri* species is consistent with other published UK epidemiological datasets on causative microbiology (7,8,21) and temporal trends in brain abscess epidemiology (6). The dominant role of *S. intermedius* is also exemplified by case reports of brain abscesses (22–26) and noted to be a frequent species in the *S. milleri* group isolated from central nervous system specimens (27). Another recent UK case series suggests the ‘*S. anginosus* group’ is predominant, but does not provide a breakdown of the *S. milleri* group by species (7). It is unclear how important speciation within the *S milleri* group is in clinical practice, given the entire group tends to be penicillin sensitive. *S. intermedius* may be dominant in brain abscesses as a result of its ecological niche in dental, sinus and ear infections (28). Its particular tendency to form abscesses may relate to the presence of specific virulence factors, including enzymes that digest host tissue (such as sialidase, hyaluronidase, and human-specific cytolysin) (29), and factors that enhance binding to fibronectin and laminin in the extra-cellular matrix (30). This is in keeping with the significantly larger size of abscesses we found in association with these organisms.

Another striking finding from the microbiological data is the relative lack of staphylococcal infections. A recent systematic review reported staphylococci as still causing approximately 20% of brain abscesses (6), but S.aureus was found in <10% of cases in another recent UK series (7), and we identified only one case caused by *S. aureus*, consistent with an ongoing downward trend. The small number of anaerobic infections in our cohort is consistent with previous literature (6,9,31). This may be partly reflective of the difficulty in culturing anaerobes, although the accumulation of more robust diagnostic microbiological data may call into question the role of anaerobes in intracranial abscesses.

### Antibiotic therapy

Our data demonstrate that ceftriaxone with metronidazole currently remains a safe empiric antimicrobial regimen in our setting (32). Narrower spectrum agents could be considered for the treatment of fully sensitive streptococci when supported by laboratory data but may be less easy to administer in one or two daily doses. Furthermore, culture-based methods may not accurately reflect the entire spectrum of organisms present, as evidence emerges for a complete microbiome within abscesses (33). A careful risk/benefit analysis is needed for inclusion of metronidazole on empiric grounds: an anaerobic contribution may be present even when not confirmed by laboratory testing, but addition of metronidazole can cause nausea, and prolonged therapy is associated with a risk of neuropathy.

Four to six weeks of therapy is typically suggested for intravenous ceftriaxone (5), but given the ongoing international drive to restrict and reduce the duration and use of broad spectrum antibiotics (34), it is important to consider whether typical prolonged parenteral antimicrobial regimens used in brain abscesses could potentially be restricted without loss of clinical efficacy, as is the case for other conditions, such as intra-abdominal infections (35), endocarditis (15) and osteomyelitis (14). Large randomised clinical trials with prospective follow-up are ideally needed to address the question about optimum route and duration of therapy, but in practice are difficult to conduct for a rare condition. The lack of guidance available on the choice, course and use of oral antibiotics (10,13) is reflected in our findings of a wide range of choices for follow-on antibiotics and their duration.

### Outcomes

Despite prompt surgical drainage and prolonged courses of antibiotic therapy, the mortality rate of this condition remains high, particularly in the context of frailty (in this setting, associated with immunocompromise, cardiac anomalies and older age). There was no increased mortality among patients in whom we did not secure a microbiological diagnosis, suggesting that empiric therapy is also covering the underlying organisms present in this group. Five deaths occurred following completion of intravenous antibiotics in the group receiving no follow-on antibiotics; however, only one of these deaths was definitively attributable to brain abscess, while the other four were associated with significant co-morbidities (all have cardiac conditions and two were significantly immunosuppressed).

### Limitations

Our data rely upon the accuracy of diagnostic coding: just as some non-brain abscess cases were erroneously coded as G06.0, some genuine brain abscess cases may have been coded with a different ICD code. This relatively low incidence rate highlights the difficulties of performing interventional trials to examine conclusions further, and any such trial would need to be coordinated across multiple centres.

Our findings apply to a specific setting in the UK and should be extrapolated with caution. We recognise that the tertiary nature of our hospital may also bias the case-mix, either as a result of over-representing complex cases that need specialist multi-disciplinary management, and/or by the exclusion of certain groups – including those who are too unwell to transfer (with high mortality) and those with simple/limited disease who are managed in local hospitals without the need for referral to a specialist centre. We restricted the remit of our audit to adults, and we are therefore unable to extend our conclusions to brain abscesses in the paediatric population. Whilst our retrospective, observational approach adds important information that may help to underpin management decisions, there is a clear need for large prospective randomised controlled trials to determine optimum choice, route and duration of therapy.

## Conclusion

From this study of the diagnosis and management of brain abscesses, we have provided contemporaneous UK epidemiological data in the context of limited published evidence. The predominant pathogen currently identified in this setting is penicillin-sensitive *S. intermedius*, but an emerging literature suggests a more complex microbiome. Multi-centre randomised prospective studies may be required to provide evidence underpinning the optimum choice, route and duration of antibiotic therapy.

## Data Availability

All data referred to in this study are included in the paper or in the supplementary data files

https://doi.org/10.6084/m9.figshare.11662950.v1

## SUPPLEMENTARY DATA

All data generated or analysed during this study are included in this published article, and its Supplementary Information files, which are accessible on-line at Figshare. Prior to publication, this is shared through a private link: https://figshare.com/s/dc1f0bb68ba2a657da81.

On acceptance for publication, this will be converted to a permanent open access DOI.

**Metadata table: xls spreadsheet containing full dataset of clinical, radiological and imaging data presented in this study**.

**STROBE statement (Strengthening the Reporting of Observational Studies in Epidemiology)**

**Suppl Figure 1: CONSORT diagram for identification of a cohort of adults with brain abscesses from a tertiary referral hospital in the UK**.

**Suppl Fig 2: Duration of antibiotic therapy for adults treated for bacterial brain abscess**

**Suppl Table 1: Association between *S. milleri* infection and other patient characteristics in a cohort of 39 adults with positive microbiology diagnosis of bacterial brain abscess**

**Suppl Table 2: Number of surgical interventions undertaken for 47 adults presenting with brain abscess**

